# Epigenome-wide association study of household air pollution exposure in an area with high lung cancer incidence

**DOI:** 10.1101/2025.04.03.25325041

**Authors:** Mohammad L Rahman, Lützen Portengen, Batel Blechter, Charles E. Breeze, Jason Y.Y. Wong, Wei Hu, George S. Downward, Yongliang Zhang, Andres Cardenas, Bou Ning, Jihua Li, Kaiyun Yang, H. Dean Hosgood, Debra T. Silverman, Nathaniel Rothman, Yunchao Huang, Roel Vermeulen, Qing Lan

**Author notes:** These authors co-supervised the work (Yunchao Huang, Roel Vermeulen, Qing Lan). To whom correspondence should be addressed: **Mohammad L Rahman,** MD, ScD, MPH Occupational & Environmental Epidemiology Branch, Division of Cancer Epidemiology and Genetics, National Cancer Institute, NIH, 9609 Medical Center Drive, Rm. 6E132, Rockville, MD 20850, USA. 9.

## Abstract

**Background:** Lung cancer incidence among never-smoking women in Xuanwei, China, ranks among the highest worldwide and is largely attributed to household air pollution (HAP) from smoky (bituminous) coal combustion, with early-life exposures possibly playing a critical role. We conducted an epigenome-wide DNA methylation (DNAm) analysis across multiple exposure windows to elucidate molecular mechanisms.

**Methods:** Leukocyte DNAm was measured in 106 never-smoking women (23 with repeated measurements). Fuel use was obtained through questionnaires, and extensive personal and environmental monitoring was conducted. Validated exposure models estimated 43 HAP constituents, primarily polycyclic aromatic hydrocarbons (PAHs), across childhood, current, and cumulative exposure windows. Hierarchical clustering derived exposure clusters. We used generalized estimating equations to identify CpG sites associated with HAP exposure and PAH clusters, including 5-methylchrysene, a methylated PAH previously linked to lung cancer.

**Results:** We identified several differentially methylated CpG sites, predominantly hypomethylated with HAP exposure. Although some DNAm signatures overlapping with smoking (cg05575921; *AHRR*) were observed, most changes were distinct. A life-course assessment indicated persistent epigenetic variations across childhood and cumulative exposures, suggesting that early-life exposures may have lasting effects at certain sites (*SLC43A2*). Within the PAH clusters, 5-methylchrysene appears to be a significant contributor to DNAm variations. Top CpG sites were linked to immune regulation, cell growth and proliferation, and molecular mechanisms of cancer, including lung cancer.

**Conclusions:** Our findings provide novel insights into HAP-induced DNAm changes and their potential health effects. Future studies with larger sample sizes, and diverse coal use settings are needed to validate and extend these findings.

## 1. INTRODUCTION

Household air pollution (HAP) poses a significant global health challenge, impacting approximately half of the world’s population, and is considered the leading environmental cause of death ^1^. Its adverse effects are predominantly observed in low and middle-income countries, particularly among women and young children ^2,3^. HAP is comprised of particulate matter (PM), volatile organic compounds, and polycyclic aromatic hydrocarbons (PAHs), among others, which are generated from incomplete combustion of solid fuels such as coal, biomass, and wood. HAP exposure has been associated with many adverse health outcomes, including respiratory and cardiovascular diseases, and cancer.

In Mainland China alone, it is estimated that over 450 million individuals still rely on solid fuels for domestic cooking and heating ^2,4^. Regions such as Xuanwei and Fuyuan counties in southwest China have exceptionally high lung cancer incidence and mortality rates among never-smoking women ^3-5^, which is largely attributed to toxic emissions from household coal combustion. For instance, we previously reported that using smoky (bituminous) coal is associated with a 100-fold increase of lung cancer risk compared to smokeless (anthracite) coal ^3,6^. Later, we found that cumulative exposure to PAHs, including 5-methylchrysene (5-MC), a highly carcinogenic PAH, was strongly associated with increased lung cancer risk in Xuanwei ^7^. Lung cancer in the Xuanwei region tends to develop at a relatively younger age, with onset occurring more than a decade earlier than the peak age observed in other parts of China ^8^.

Historically, households in this region relied on smoky coal for cooking and heating, with many continuing this practice into adulthood, resulting in a consistent lifetime exposure pattern. Using a life-course approach, we previously established an association between PAH exposure and lung cancer risk among smoky coal users in Xuanwei, emphasizing the critical role of early-life exposures ^9^. Although the underlying molecular mechanisms remain unclear, HAP exposure from smoky coal combustion, particularly during early life, may drive epigenetic alterations that increase lung cancer susceptibility in this population.

Epigenetic alterations, particularly DNA methylation (DNAm), are sensitive to environmental exposures and are implicated in mediating the relationship between environmental factors and chronic diseases ^10^. To our knowledge, no population-based epigenome-wide association study (EWAS) has examined the effects of HAP exposure from solid fuel use. We recently reported that PAH exposure, including 5-MC, was significantly associated with epigenetic age acceleration, as measured by the GrimAge clock in this population ^11^. The GrimAge clock is a DNAm-based marker of biological aging that can predict mortality and morbidity of many diseases, including cancer ^12^. We subsequently found that GrimAge acceleration was significantly associated with an increased future risk of lung cancer among never-smokers in Shanghai, China ^13^. These findings lay the groundwork for understanding HAP-induced epigenetic modifications and their potential impacts on lung cancer risk.

In this study, we investigated the association between HAP exposure from smoky coal combustion and DNAm patterns in peripheral blood leukocyte among women who never smoked. Given that both smoky coal combustion and tobacco smoke contain PAHs implicated in lung cancer, we implemented a two-stage analysis. First, we targeted CpG sites previously associated with smoking to determine whether HAP exposure among never smokers induces similar epigenetic alterations as smoking. Next, we conducted an agnostic epigenome-wide analysis to identify novel DNAm changes linked to HAP exposure. Additionally, we examined the influence of exposure windows—childhood, current, and cumulative lifetime exposures—on DNAm patterns to determine how the timing of HAP exposure may shape epigenetic alterations in this population.

## 2. METHODS

### 2.1. Study population

The study population has been previously described in detail ^2,9^. Briefly, a total of 163 healthy women who never smoked were sampled from 30 villages in Xuanwei and Fuyuan counties, located in the Yunnan province of China. Recruitment occurred during two distinct time intervals: August 2008 to February 2009, and March to June 2009. During the initial collection period, all 30 villages were visited, and a total of 148 participants were recruited. In the subsequent period, 16 villages were revisited, representing the overall population, and 53 of the original participants were reevaluated, while an additional 15 new participants were recruited. From each village, up to five households were chosen, and one nonsmoking woman aged 20 to 80 from each home was recruited. Homes were prioritized if they were at least ten years old and had not replaced their solid fuel stoves in the past five years, reflecting the region’s historical patterns of stove and fuel use.

All participants provided written informed consent, and the study received approval from the National Cancer Institute Special Studies Institutional Review Board.

### 2.2. Solid fuel use

Trained interviewers conducted in-person interviews to gather demographic, anthropometric, and household information. A questionnaire was utilized to document women’s household activities during the measurement period and captured details on the type of stove and ventilation system in the household, cooking practices, heating methods, the specific type of coal mine that supplied the household fuel, and fuel usage. The fuel used during the measurement period was categorized as smokeless coal, smoky coal, and wood or plant material. In comparison, childhood fuel usage was categorized as smokeless coal, smoky coal, wood, or mixed fuel (a combination of coal, wood, and plant material).

### 2.3. Estimation of household air pollution constituents

Exposure assessment has been described in detail previously ^9,14,15^. Personal and indoor measurements were collected over two sequential 24-h periods, with approximately half of the subjects revisited in a subsequent season to capture seasonal variability. Samples of particle bound PAHs, including methylated PAHs, PM_2.5_, black carbon (BC) were collected on 37 mm Teflon filters. Indoor measurements of NO_2_ and SO_2_ were collected using passively diffusing filters (Ogawa). Additionally, self-reported data on stove use, fuel type, and the coal source were recorded for each participant. Supervised stepwise predictive linear mixed-effect models were then employed to estimate annual average exposures for each pollutant, using self-reported stove and fuel histories ^7,11,15,16^. In these models, village and individual were treated as random effects, and fixed effects included fuel usage, stove design, room volume, and the season of measurement. Model performance was evaluated using the Akaike Information Criterion (AIC) and the ratio of the variance of predicted to observed values (R^2^).

### 2.4. Derivation of cluster prototypes

Exposure clusters were constructed using hierarchical cluster analysis, accounting for a strong correlation among individual pollutants, as explained previously ^7,9^. Clusters were created for various exposure time points, including current (during the measurement period), childhood (age 0-18 years), and cumulative exposures (lifetime), and were represented as the first principal component, mean-centered and scaled, of each cluster. A list of all cluster prototypes for all three exposure time-points has been presented in the Table S1. In this study, we focused our analysis on the largest PAH clusters derived for the current (31 PAH constituents, PAH31), childhood (33 PAH constituents, PAH33), and cumulative (36 PAH constituents, PAH36) exposures, as these clusters showed the most consistent associations with lung cancer ^9^ and epigenetic age acceleration ^11^ in this population. In addition, we separately examined 5-MC, a highly carcinogenic PAH, that emerged as a key component within the broader PAH clusters, in relation to lung cancer ^7^ and accelerated aging ^11^.

### 2.5. DNA extraction and methylation measurements

DNA was extracted from leukocytes obtained from peripheral blood collected during the second visit. Standard procedures were followed to extract DNA from whole blood. Bisulfite conversion was applied to the DNA samples, which were then randomized across the chip. Methylation levels at over 485,000 CpG sites were quantified using the Infinium HumanMethylation450 BeadChip assay, following the manufacturer’s protocol (San Diego, CA, USA).

We processed raw DNAm image files using R statistical software (www.r-project.org/). We used several Bioconductor packages including the ChAMP pipeline for quality control using default parameters. No individual samples were removed based on the detection p-value <0.10 while 9,927 probes were removed based on the same threshold. In addition, 506 probes with bead count <3 in at least 5% of samples, 2,817 non-CpG probes, 57,037 SNP-associated probes, and 11 multi-hit probes were removed ^17^. Mixture Quantile dilation (BMIQ) was used to normalize beta values. A total of 415,214 CpG sites passed quality controls and were subsequently used in statistical analyses. Finally, we corrected for sample plate, array, and slide using ComBat ^18^. DNA-methylation level at each CpG locus was measured as M-values (logit2-transformation of β-values, the per-site methylation fraction).

We visualized density distributions for samples at all processing steps (Figure S1). For each CpG site, DNAm is reported as β-values, corresponding to an interval-scaled quantity between zero and one, which is interpreted as the fraction of DNA molecules whose target CpG is methylated. We used the Houseman’s method for inference of cell-type composition, to estimate cell-type proportions for CD8+T cells, CD4+T cells, NK, B-cells, monocytes, and neutrophils ^19^.

### 2.6. Statistical Analysis

#### 2.6.1 Investigating associations with previously identified smoking-related CpG sites

We initially restricted our analyses to 2,671 CpG sites previously reported as differentially methylated between never and current or former smokers ^20^. After probe filtering, 2,476 smoking-related CpG sites were retained. Given prior evidence suggesting that PAHs play a significant role in lung cancer development in this population ^7,9^, our goal was to identify both shared and distinct DNAm patterns associated with PAH exposure from smoky coal combustion and cigarette smoking.

We used generalized estimation equation (GEE) models for each CpG sites (M-values), accounting for repeated measures and adjusting for age, body mass index (BMI; kg/m^2^), county of residence (Xuanwei or Fuyuan), education (no education attended elementary school, graduated elementary school, attended middle school or higher), and socioeconomic status (SES; no luxury items and at least one luxury item such as a bicycle, sewing machine, radio, watch, phone, motorcycle, TV set or tractor), and estimated cell type proportions. Analyses were conducted using smokeless coal as the reference group for smoky coal use. Additionally, we examined these CpGs in relation to clusters of PAH constituents across childhood, current, and cumulative lifetime exposure windows, as well as 5-MC. Statistical significance for the targeted analysis was defined using a Bonferroni-adjusted threshold of *p* < 0.05/2,476 (2.20 × 10 ^5^).

Additionally, we calculated a pack-years methylation score as previously described ^21^. This score, derived from smoking-related signatures identified in large-scale EWAS meta-analyses ^20^, provides a composite measure of smoking-induced DNAm changes and correlates with smoking-related gene expression alterations ^21^. To evaluate the extent to which this score is correlated with PAH exposures from smoky coal combustion, independent of smoking, we calculated Spearman correlation coefficients across exposure windows.

#### 2.6.2 Agnostic epigenome-wide association analysis

For computational efficiency, we initially screened epigenome-wide associations using robust linear regression models combined with a lenient Benjamini-Hochberg false discovery rate (FDR) threshold of <0.05. CpG sites meeting this threshold were subsequently reanalyzed using GEE models to account for repeated measurements. Ultimately, epigenome-wide significance was determined using a Bonferroni-adjusted threshold of p < 0.05/415,214 (1.20 × 10□□). Analyses were performed for smoky coal use, PAH clusters, and 5-MC across different exposure windows.

#### 2.6.3 Integrative bioinformatics analysis

EWAS hits for smoky coal use (1894 CpG sites at FDR<0.05) were further analyzed for integrative bioinformatics analysis to derive biological relevance of our findings. These analyses included cell-type specific functional overlap analysis using eFORGE (version 2.0, https://eforge.altiusinstitute.org/)^22,23^ and ingenuity pathway analysis (IPA, version 01-23-01, Qiagen) ^24,25^.

#### 2.6.3.1 Cell-type specific functional overlap analysis using eFORGE

eFORGE analysis evaluates whether differentially methylated CpG sites are enriched for putative functional elements relative to matched background sites. By assessing enrichment on a per-cell-type basis, eFORGE identifies tissue-specific signals, revealing potential sites of action underlying the EWAS signature. The analysis examined enrichment across DNase I hotspots, Hidden Markov Model (HMM) chromatin states, and histone mark broad peaks from the Roadmap Epigenomics Consortium [24].

#### 2.6.3.2 Ingenuity Pathway Analysis (IPA)

IPA analysis included canonical pathway analysis, molecular network analysis, and upstream regulator analysis. The canonical pathway analysis is conducted based on the IPA knowledge base, which is the largest curated database of previously published findings on mammalian biology. Molecular networks in IPA examines interactions between molecules within the dataset and their relevance to various diseases or biological functions. Networks are generated based on curated molecular interactions, with networks ranked by a score, where higher scores indicates greater confidence that the molecules within the network are functionally related in the context of the studied exposure or condition ^26^.

IPA calculates p-values using a right-tailed Fisher’s exact test to evaluate the statistical significance of overlap between input genes and genes in a given pathway, with a significance threshold set at FDR < 0.05. IPA also calculates ‘activation Z-scores’ to predict the activation or inhibition state of pathways based on the direction of expression changes in the input data. We made a simplified assumption that hypomethylation corresponds to increased gene expression, recognizing that this relationship may not always hold true across all genomic contexts. A positive ‘activation Z-score’ suggested activation, while a negative score indicated inhibition, with |Z| > 2 considered significant. The algorithms used for calculating p-values and ‘activation Z-scores’ have been described previously ^24^.

## 3. RESULTS

### 3.1. Study population characteristics

Table 1 presents the demographic details of the study population. DNAm data were obtained for 106 women in the study, with 91 participants sampled during Visit 1 and an additional 15 enrolled at Visit 2. Additionally, 23 participants from Visit 1 had repeat measurements taken during Visit 2. The average age was 56.2 years (standard deviation [SD] = 15.0), and the mean BMI was 22.0 kg/m^2^ (SD = 3.5). Half of the participants were from Xuanwei County (50.0%), and 50.0% had at least one luxury item in the household. Most women used smoky coal during the measurement period (77.4%) and lacked formal education (67.9%).

**Table 1.** Demographic characteristics and the distribution of indoor air pollution among 106 never-smoking women within the Xuanwei Exposure Assessment Study.

| Characteristics | n (%) |
| --- | --- |
| Age in years; mean ± SD | 56.24 ± 15.03 |
| BMI; mean ± SD | 21.99 ± 3.46 |
| Education |  |
| No formal education | 72 (67.9) |
| Attended elementary school | 17 (16.0) |
| Graduated elementary school or higher | 17 (16.0) |
| Socioeconomic status |  |
| No luxury items in household | 53 (50.0) |
| At least one luxury item in household | 53 (50.0) |
| Study visit |  |
| Visit 1 | 91 (85.8) |
| Visit 2 | 15 (14.2) |
| County |  |
| Fuyuan | 53 (50.0) |
| Xuanwei | 53 (50.0) |
| Current fuel type |  |
| Smokeless | 13 (12.3) |
| Smoky | 82 (77.4) |
| Wood or Plant | 11 (10.4) |
| Exposure to PAH, ng/m<sup>3</sup>-year |  |
| Childhood PAH33; mean ±SD | 0.13 ± 1.00 |
| Current PAH31; mean ±SD | 0.21 ± 0.98 |
| Cumulative PAH36; mean ±SD | 0.14 ± 1.00 |
| Exposure to 5-MC, ng/m<sup>3</sup>-year |  |
| Childhood 5-MC; mean ±SD | 92.03 ± 50.20 |
| Current 5-MC; mean ±SD | 8.13 ± 4.14 |
| Cumulative 5-MC; mean ±SD | 265.98 ± 148.59 |

Abbreviations: SD, standard deviation; BMI, body mass index; PAH, polycyclic aromatic hydrocarbon; 5-MC, 5-methylcrysene.
A total of 23 participants had one repeat measurements.
PAH and 5-MC exposure data were missing in 3 participants.

### 3.2. Correlations of HAP constituents across exposure windows

The correlations between PAH exposure measures across exposure windows are presented in Table S2. Childhood and cumulative exposures exhibited strong correlations for both PAH clusters (ρ = 0.77, P < 2.2 × 10^−16^) and 5-MC (ρ = 0.77, P < 2.2 × 10^−16^), while current exposures showed moderate correlations with childhood and cumulative exposures (ρ = 0.42 –0.65). When comparing PAH and 5-MC, cumulative exposures were highly correlated (ρ = 0.93, P < 2.2 × 10^−16^), with similarly strong correlations observed for childhood exposures (ρ = 0.88, P <2.2×10^−16^).

### 3.3. Epigenetic signature of HAP exposure: Links to smoking-related DNAm patterns

#### 3.3.1. Associations between smoky coal use and smoking-related CpG sites

HAP exposure from smoky coal combustion was associated with differential methylation of 28 smoking-related CpG sites compared to smokeless coal combustion (FDR<0.05 (Figure 1a and 1c, Supplementary table 2). Of these, nine CpG sites surpassed the Bonferroni threshold (P < 2.02 × 10□□). All but one (cg16672562, *HIF3A)* exhibited significant hypomethylation with smoky coal combustion, including cg04907244 and cg22407942 (*SNORD93*); cg15845821 and cg20249566 (*NWD1*), cg11152412 (*EDC3*), cg14817490 (*AHRR*), cg24981097 (*STK24*), cg01962471 (*ZDHHC14*). Of these, the first five sites – annotated to *SNORD93* (2 sites), *NWD1* (2 sites), and *EDC3* – had prior associations with chronic obstructive pulmonary disease (COPD) ^27^, an independent risk factor for lung cancer in both smokers and never smokers ^28,29^. The strongest association was observed for cg04907244 (*SNORD93;* P = 4.5 × 10□¹□), with a 66% of lower average methylation M-value for smoky coal combustion compared to smokeless coal use.

**Figure 1.**
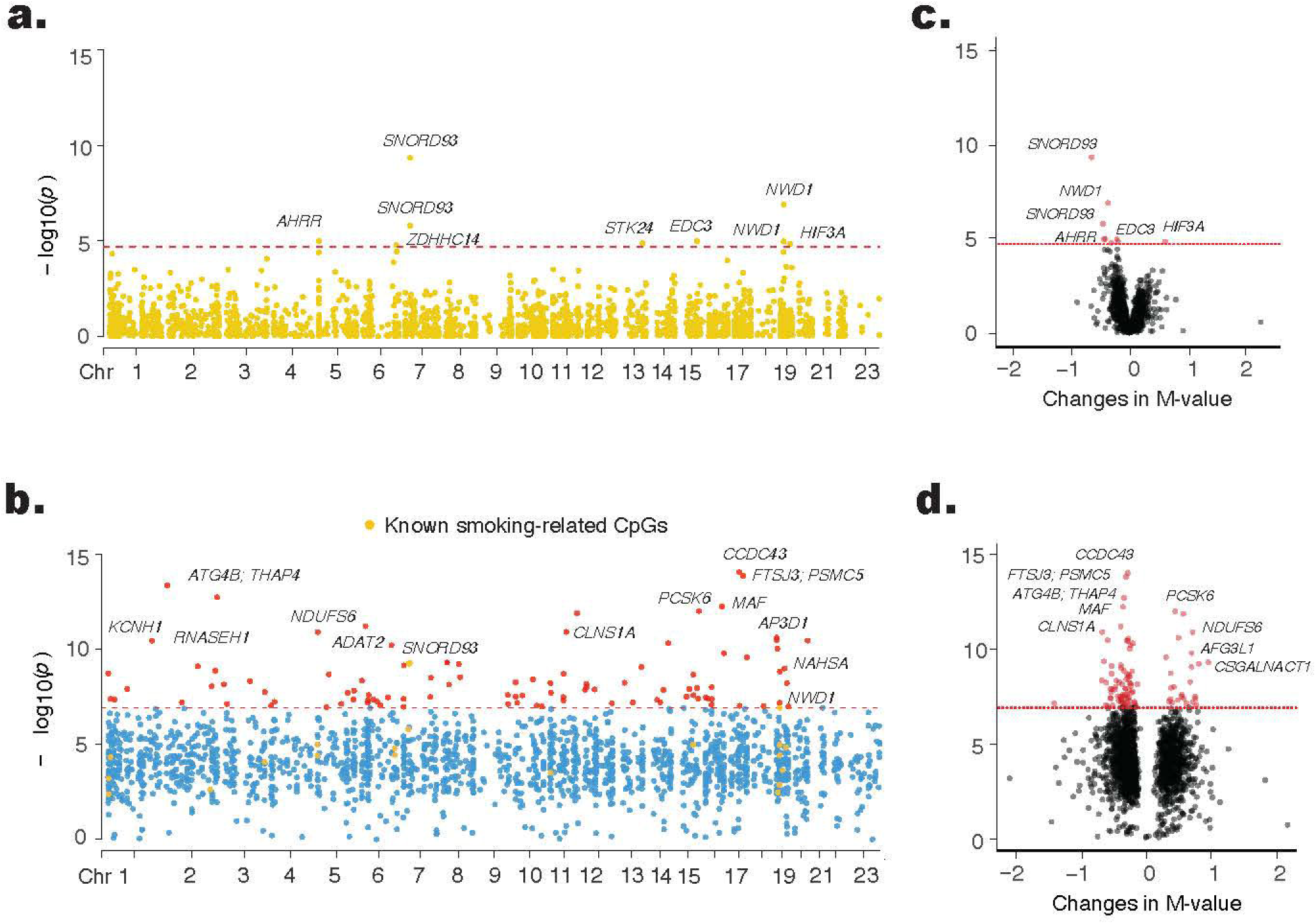
Association between HAP exposure from smoky coal combustion and leukocyte DNA methylation. Generalized estimating equation models were used to account for repeated measurements, adjusting for age, BMI, education, SES, county, and estimated cell type proportions. Manhattan plots depict associations between CpG sites and exposure to smoky coal combustion. **Panel (a)** shows results for 2,476 smoking-related CpG sites, while **panel (b)** presents epigenome-wide associations for 415,249 CpG sites. The x-axis represents genomic locations by chromosome number, and the y-axis displays the −log_10_ (*p*-value). The red horizontal dotted line indicates the Bonferroni-corrected significance threshold of *p* = 0.05/2,476 (2.01 × 10□□) for the targeted analysis in panel (a) and *p* = 0.05/415,249 (1.20 × 10□□) for the agnostic epigenome-wide analysis in panel (c). CpG sites exceeding the significance threshold in panel (c) are highlighted in red, except for known smoking-related CpG sites, which are colored in gold. Volcano plots illustrate the effect estimates (x-axis) and −log_10_ (*p*-value) (y-axis) for the association between smoky coal exposure and leukocyte DNA methylation. **Panel (c)** focuses on 2,476 smoking-related CpG sites, and panel (**d**) presents epigenome-wide associations for 415,249 CpG sites. The red horizontal dotted line indicates the Bonferroni-corrected significance thresholds of *p* = 2.01 × 10 and p = 1.20 × 10□□, respectively. CpG sites exceeding these thresholds are highlighted in red.

#### 3.3.2. Associations between PAH clusters across exposure windows and smoking-related CpG sites

Higher childhood exposure to the PAH cluster was associated with significant hypomethylation at two CpG sites (cg09219877, *DNASE1L2*; cg09938479), with cg09938479 along with two additional CpG sites (cg01076459, *RAP1GAP2*; cg11152384), also showing significant hypomethylation with cumulative PAH exposure at the Bonferroni threshold (P < 2.02 × 10□□; (Figure 2, Supplementary Table 3). Higher current exposure to the PAH cluster was associated with significant hypomethylation at four CpG sites (cg12147622, cg01127300, cg26657675, cg16474118). Notably, the first three of these sites have been previously linked to COPD ^27^.

**Figure 2.**
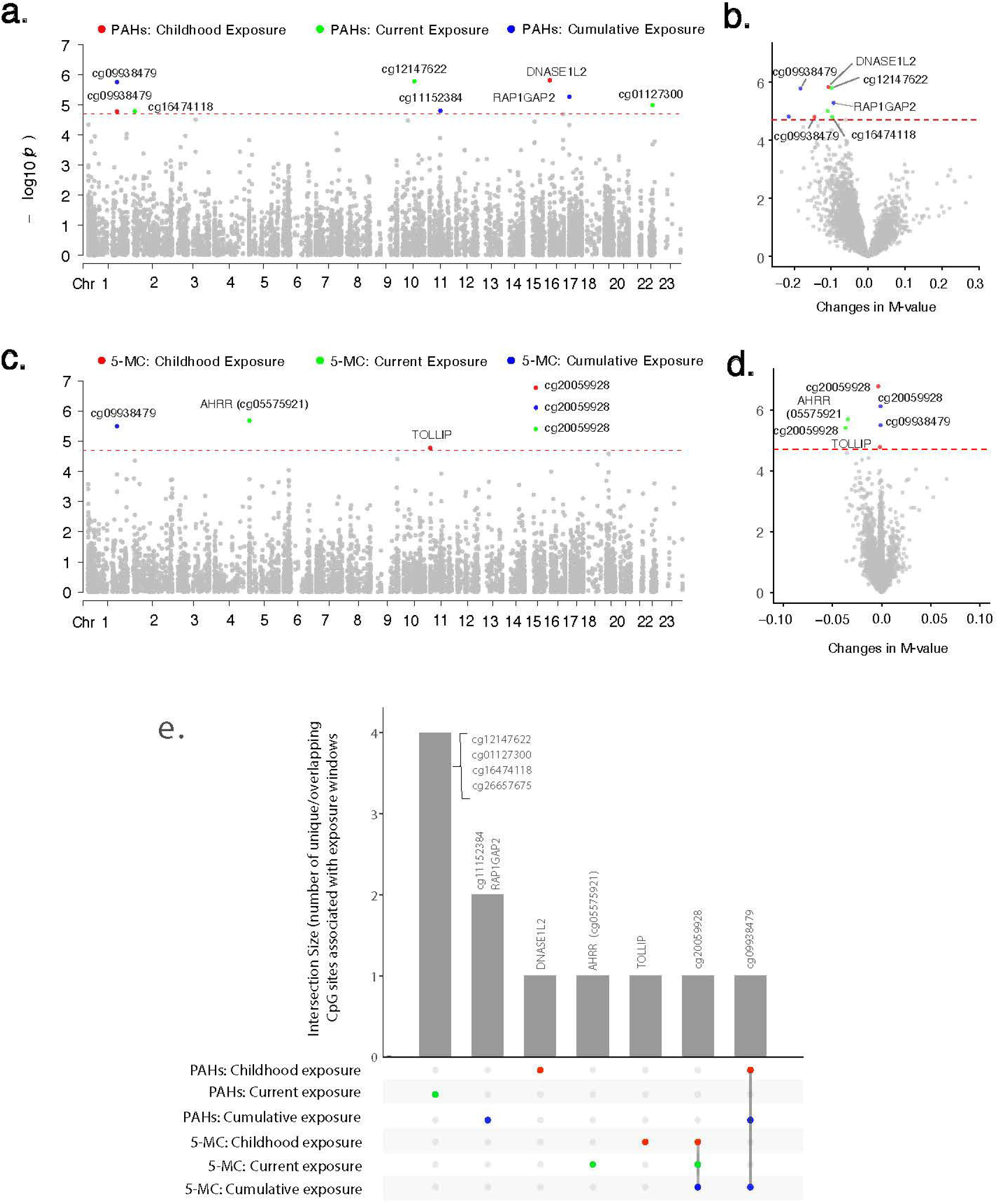
Associations between PAH clusters, 5-MC, and leukocyte DNA methylation at 2,476 smoking-related CpG sites across childhood, current, and cumulative lifetime exposure windows. Generalized estimating equation models were used to account for repeated measurements, adjusting for age, BMI, education, SES, county, and estimated cell type proportions. Manhattan plots depict associations between childhood, current, and cumulative lifetime exposures to household smoky coal combustion constituents and 2,476 smoking-related CpG sites. Panel (**a)** shows results for PAH clusters, while panel (**c**) presents results for 5-MC. The x-axis represents the genomic locations of CpG sites by chromosome number, and the y-axis shows the −log_10_ (*p*-value). Each dot represents a smoking-related CpG site. The red horizontal dotted line indicates the Bonferroni-corrected significance threshold of *p* = 2.01 × 10□□. CpG sites exceeding the significance threshold are highlighted in red for childhood exposure, turquoise for current exposure, and blue for cumulative exposure, while those below the threshold are shown in gray. Volcano plots illustrate associations between childhood, current, and cumulative lifetime exposures to household smoky coal combustion constituents and 2,476 smoking-related CpG sites. Panel (**b)** shows results for PAH clusters, while panel (**d**) presents results for 5-MC. The x-axis represents effect estimates as changes in methylation M-values, and the y-axis shows the −log_10_ (*p*-value). The red horizontal dotted line indicates the Bonferroni-corrected significance threshold of *p* = 2.01 × 10□□. CpG sites exceeding the significance threshold are highlighted in red for childhood exposure, turquoise for current exposure, and blue for cumulative exposure, while those below the threshold are shown in gray. Panel (**e**) presents UpSet plot showing the intersection of top smoking-related CpG sites associated with PAH clusters and 5-MC across childhood, current, and cumulative lifetime exposures. The combination matrix at the bottom (x-axis) shows overlapping CpG sites across different exposure windows (red = childhood, turquoise = current, blue = cumulative lifetime), while the bars above indicate the number of CpG sites in each intersection (y-axis).

#### 3.3.3. Associations between 5-MC across exposure windows and smoking-related CpG sites

Higher childhood exposure to 5-MC was associated with significant hypomethylation at two CpG sites (cg20059928 and cg10696445, *TOLLIP*) at P < 2.02 × 10 (Figure 2c and 2d, Supplementary Table 3). Notably, cg20059928, previously linked to COPD incidence ^27^, also exhibited significant hypomethylation with higher current and cumulative exposure to 5-MC, indicating persistent changes in DNAm levels at this site.

Interestingly, higher current exposure to 5-MC was associated with significant hypomethylation of the well-known smoking-related CpG site, cg05575921, located at the *AHRR* gene. In contrast, the associations with childhood and cumulative exposure to 5-MC were weaker for this CpG site, following similar patterns observed for PAH clusters across exposure windows.

Additionally, higher cumulative exposure to 5-MC was significantly associated with hypomethylation of cg09938479, a CpG site that also exhibited similar associations with childhood and cumulative exposure to PAH clusters. This finding suggests that the effect of PAH clusters on this CpG site was likely driven by 5-MC.

#### 3.3.4. Associations between PAH exposure across exposure windows and DNAm-based smoking score

We observed modest correlations between cumulative PAH exposure (*ρ* = 0.31, P = 5.1 x 10^-4^) and the DNAm-based smoking pack-years even among women who never smoked, while correlations for childhood (*ρ* = 0.14, P = 0.11) and current exposures (*ρ* = 0.07, P = 0.42) were very weak (Table S2). Similar patterns were observed for 5-MC (*ρ* for cumulative 5-MC= 0.26, P = 3.3 x 10^-3^). These findings suggest that sustained PAH exposure from non-smoking environmental sources, such as that from smoky coal combustion, may accumulate over time and leave a detectable epigenetic signature that overlaps with smoking-related DNAm patterns, even among never-smokers.

### 3.4. Agnostic epigenome-wide associations of HAP exposure

#### 3.4.1. Associations of HAP exposure from smoky coal combustion

HAP exposure from smoky coal combustion was associated with differential methylation at 1,895 CpG sites (FDR<0.05) in robust linear regression models, including 20 smoking-related sites. These associations were further evaluated using GEE models, which accounted for repeated measurements, with effect sizes correlated >0.93 between the two models. The GEE model identified 100 CpG sites significantly associated with HAP exposure at the epigenome-wide significance threshold (P < 1.20 × 10□□; Figure 1b and 1d, Supplementary Table 4). Most of these sites (75%) exhibited hypomethylation in response to HAP exposure, with the top-ranked site, cg11234017 (*CCDC43*), reaching P = 9.21 × 10□¹□. Notably, two CpG sites, cg04907244 (*SNORD93*) and cg27139419 (*IGF1R*), including one previously known smoking-related site (cg04907244, *SNORD93*), had prior associations with COPD incidence ^27^.

#### 3.4.2. Associations of PAH clusters across exposure windows

Epigenome-wide analysis revealed that DNAm patterns associated with PAH clusters were largely distinct from cigarette smoking. Childhood exposure to the PAH cluster was associated with six CpG sites at P < 1.20 × 10□□, four of which were hypomethylated (cg04320476, cg12564034, and cg15090217, all annotated to *SLC43A2*, and cg17105014, annotated to *GYPC*), while two were hypermethylated (cg19326543, *LIMS2* and cg23109721). Two of these CpG sites (cg04320476 and cg15090217, annotated to *SLC43A2*) were also associated with cumulative PAH exposure. The strongest association was observed for cg04320476, where each SD increase in exposure corresponded to a 47.9% (P = 4.0 × 10 ¹³) and 47.0% (P = 4.9 × 10□¹□) reduction in methylation M-values for childhood and cumulative PAH exposure, respectively. In contrast, current PAH exposure was associated with a weaker 20% reduction (P = 6.9 × 10 ³) at this site. Cumulative exposure to the PAH cluster was further associated with 17 additional CpG sites, with all but three showing hypomethylation. In general, these CpG sites exhibited similar associations with childhood PAH exposure but showed weaker associations with current exposure. Current exposure to PAH cluster was associated with five CpG sites, all but one (cg12626036) exhibiting significant hypomethylation (cg03049196, *LRDD*; cg11707352, *CITED1*; cg14788660, *FDPS*; and cg24189721).

#### 3.4.3. Associations of 5-MC across exposure windows

5-MC exhibited a similar association pattern to the PAH cluster (Figure 3, Supplementary Table 5). Specifically, higher childhood exposure to 5-MC was significantly associated with hypomethylation at three CpG sites (cg04320476, cg12564034, and cg15090217, all annotated to *SLC43A2*), which were also linked to childhood exposure to the PAH cluster. These sites exhibited similar associations with cumulative exposure to both 5-MC and PAH clusters.

**Figure 3.**
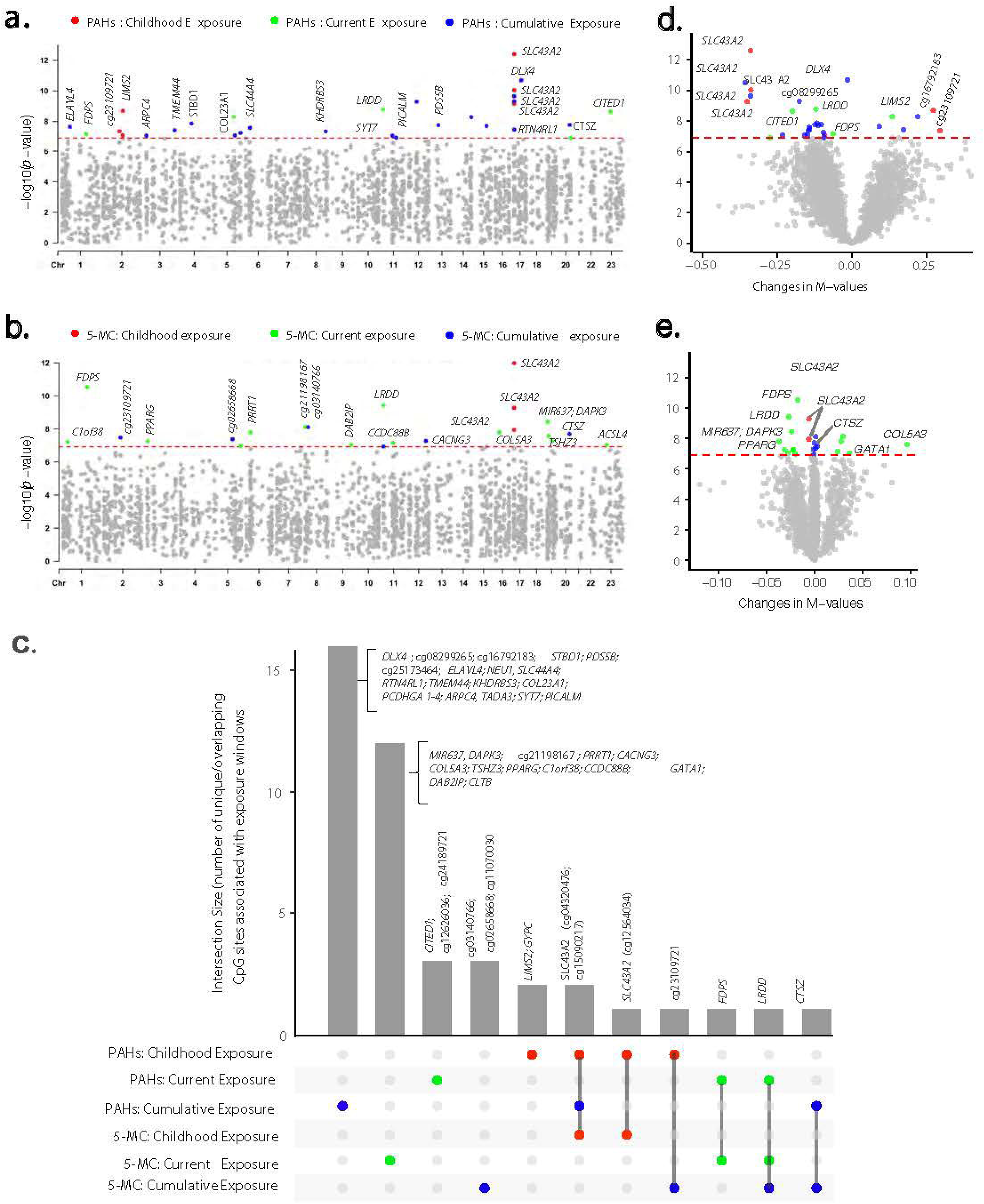
Epigenome-wide associations between PAH clusters, 5-MC, and leukocyte DNA methylation across childhood, current, and cumulative lifetime exposures. Of 415,249 CpG sites initially assessed using robust linear regression models—adjusting for age, BMI, education, SES, county, and estimated cell type proportions—785 CpG sites passed the false discovery rate (FDR < 0.05) threshold. These sites were subsequently evaluated using generalized estimating equation models, which accounted for repeated measurements and adjusted for the same set of covariates (results presented). Manhattan plots depict epigenome-wide associations for childhood, current, and cumulative lifetime exposure to household smoky coal combustion constituents. Panel (**a)** shows results for PAH clusters, while panel (**c)** presents results for 5-MC. The x-axis represents the genomic locations of CpG sites by chromosome number, and the y-axis shows the −log_10_ (*p*-value). Each dot represents a CpG site. The red horizontal dotted line indicates the Bonferroni-corrected significance threshold of *p* = 1.20 × 10 based on 415,249 CpG sites. CpG sites exceeding the threshold are highlighted in red for childhood exposure, turquoise for current exposure, and blue for cumulative exposure, while those below the threshold are shown in gray. Volcano plots illustrate epigenome-wide associations for childhood, current, and cumulative lifetime exposure to household smoky coal combustion constituents. Panel (**b)** shows results for PAH clusters, while panel (**d)** presents results for 5-MC. The x-axis represents effect estimates as changes in methylation M-values, and the y-axis shows the −log_10_ (*p*-value). The red horizontal dotted line indicates the Bonferroni-corrected significance threshold of *p* = 1.20 × 10 based on 415,249 CpG sites. CpG sites exceeding the threshold are highlighted in red for childhood exposure, turquoise for current exposure, and blue for cumulative exposure, while those below the threshold are shown in gray. Panel (**e**) presents UpSet plot showing the intersection of epigenome-wide significant hits associated with PAH clusters and 5-MC across childhood, current, and cumulative lifetime exposures. The combination matrix at the bottom (x-axis) shows overlapping CpG sites across different exposure windows (red = childhood, turquoise = current, blue = cumulative lifetime), while the bars above indicate the number of CpG sites in each intersection (y-axis).

Current exposure to 5-MC was significantly associated with 14 CpG sites, with all but five exhibiting hypomethylation in response to increasing exposure. The two top-ranked CpG sites (cg14788660, *FDPS*; cg03049196, *LRDD*) were also associated with current exposure to the PAH cluster, while cg03049196 (*LRDD*) was additionally linked to cumulative exposure to 5-MC. The *LRDD* gene, also known as P53-induced Death Domain Protein 1 (PIDD1), plays a crucial role in apoptosis and DNA damage response by mediating p53-dependent cell death pathways ^30^. In addition, cumulative exposure to 5-MC was associated with five additional CpG sites, two of which were hypomethylated (cg21120539, *CTSZ*; cg11070030), while three were hypermethylated (cg03140766; cg23109721; cg02658668) with increasing exposure. Notably, cg21120539 (*CTSZ*) was also associated with cumulative exposure to the PAH cluster, while cg23109721 was linked to childhood exposure to the PAH cluster.

Although 5-MC was not associated with any previously established smoking-related CpG sites at the epigenome-wide significance level, one known smoking-related site, cg20059928, ranked among the top five CpG sites for childhood 5-MC exposure (P = 1.68 × 10□□) and among the top 25 for cumulative 5-MC exposure (P = 7.62 × 10□□). Similarly, cg05575921 (*AHRR*) (P = 2.05 × 10□□) ranked among the top 60 CpG sites for current 5-MC exposure.

### 3.5. Findings from integrative bioinformatics analysis

#### 3.5.1. Findings from cell-type specific functional overlap analysis

The DNAm changes associated with smoky coal use were predominantly enriched in T cell regulatory elements marked by DNase I hypersensitive sites, with the strongest enrichment observed in primary T cells (E034, Figure 4). Further investigation revealed that these changes were predominantly located in promoters (H3K4me3) and enhancers (H3K4me1) in T cells. A more detailed analysis identified active transcription start sites (TssA) and enhancer chromatin states specifically in CD8 memory T cells (E048: Primary CD8 memory T cells from peripheral blood). These findings suggest a potential link between smoky coal exposure and immune dysregulation, particularly through altered leukocyte DNAm patterns predominantly affecting CD8 cytotoxic T cell. Given that cytotoxic T cells play a central role in anticancer immunity and serve as the foundation for current cancer immunotherapies ^31^, these epigenetic modifications may have implications for immune function and cancer susceptibility.

**Figure 4.**
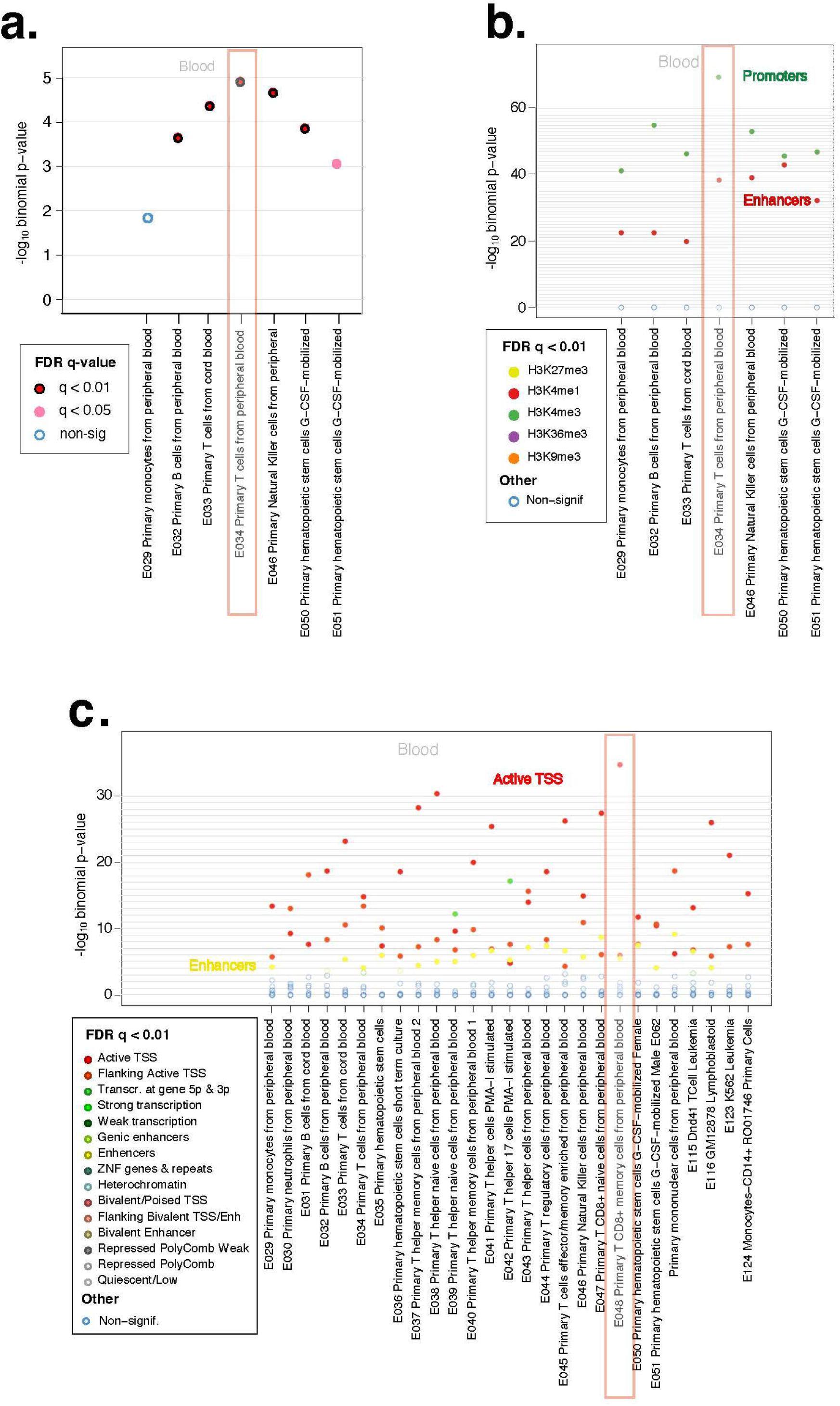
eFORGE analysis of the top 1,000 CpG sites associated with household exposure to smoky coal combustion in never-smoking women in Xuanwei, China. Full results are presented in Supplementary Materials. Panel (**a**) shows results from DNase I hypersensitive sites (DHS) analysis. The x-axis represents tissues or cell types, and the y-axis displays eFORGE enrichment (−log_10_ *p*-value) based on Roadmap Epigenomics data. The highest enrichment is observed for “E034 primary T cells from peripheral blood”, indicating that smoky coal-related DNA methylation changes predominantly occur in T cell regulatory elements marked by DNase I hotspots. Panel (**b**) shows results from histone mark analysis. The x-axis represents tissues or cell types, and the y-axis displays eFORGE enrichment (−log_10_ *p*-value) based on histone mark broad peaks from Roadmap Epigenomics data. The highest enrichment is again observed for “E034 primary T cells”, suggesting that the T cell regulatory elements affected by smoky coal exposure are primarily promoters (H3K4me3) and enhancers (H3K4me1). Panel (**c**) shows results from chromatin state analysis. The x-axis represents tissues or cell types, and the y-axis displays eFORGE enrichment (−log_10_ *p*-value) based on chromatin states from Roadmap Epigenomics data. The highest enrichment is observed for “E048 primary T CD8+ memory cells from peripheral blood”, indicating that T cell regulatory elements affected by smoky coal exposure are predominantly in regions with active transcription start sites (TssA) and enhancers in CD8+ memory T cells.

#### 3.5.2. Canonical pathway analysis

A total of 38 canonical pathways were enriched with genes annotated to smoky coal-related CpG sites, meeting the criteria of FDR < 0.05 and an absolute activation Z-score > 1.0, which indicates the predicted direction of pathway activation (Figure 5, Table S7). All but two pathways were activated in response to smoky coal combustion.

**Figure 5.**
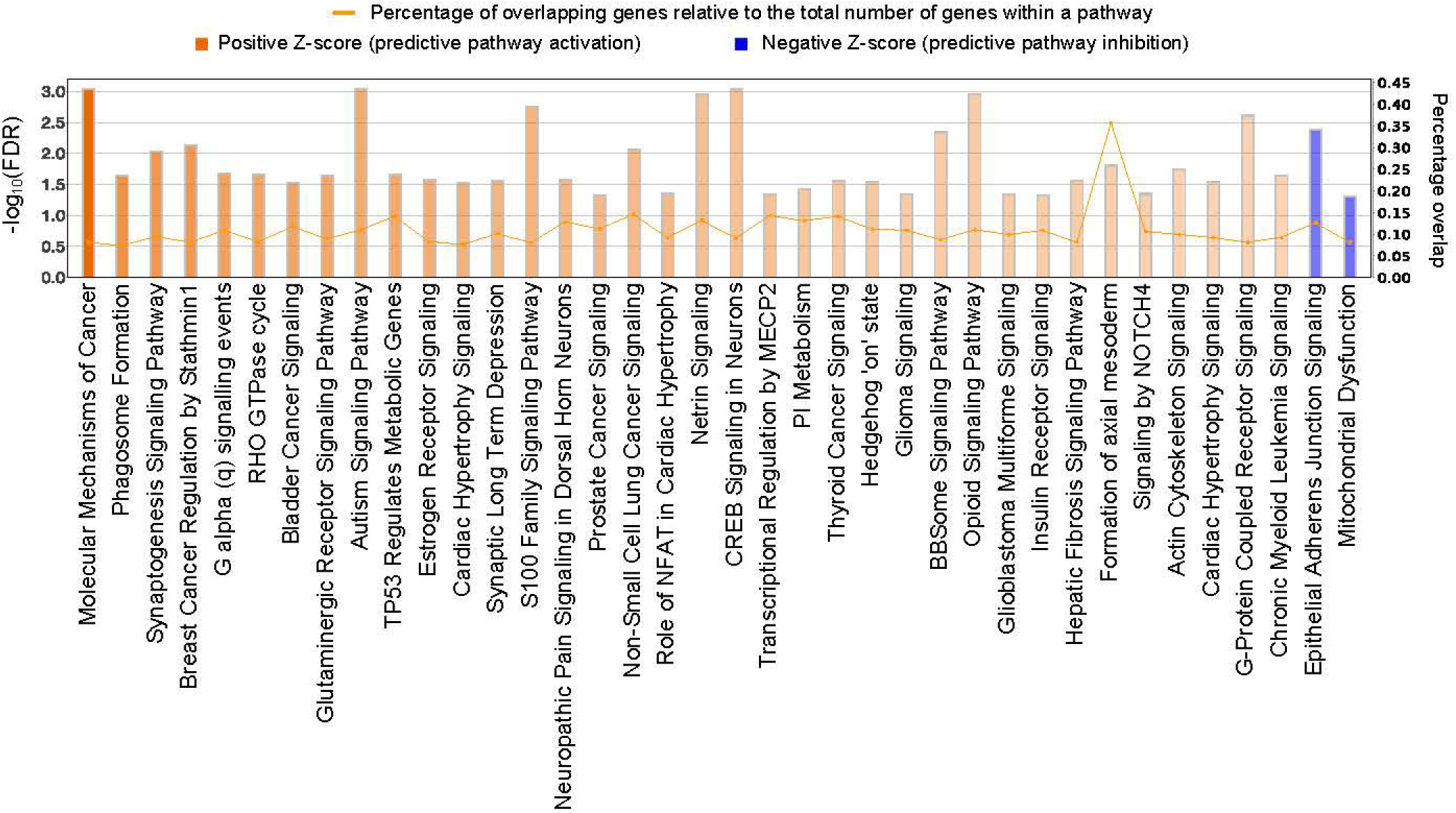
Histogram of the 38 canonical pathways enriched with CpG sites associated with HAP exposure from smoky coal combustion. The left y-axis represents the log-transformed false discovery rate (log[FDR]), while the right y-axis denotes the percentage overlap, calculated as the proportion of genes in each canonical pathway that are present in the input dataset relative to the total number of genes known to be involved in that pathway. The color gradient of the bars indicates the activation Z-score, reflecting the predicted direction of pathway regulation: orange represents pathway activation, while blue denotes predicted inhibition in response to smoky coal exposure.

The ‘molecular mechanisms of cancer’ pathway ranked highest (FDR = 9.1 x 10^-4^; activation Z-score = 4.03), reflecting significant activation in response to smoky coal use. Additionally, several cancer signaling pathways were activated, including the ‘non-small cell lung cancer signaling’ pathway (FDR = 8.5 × 10 ³; activation Z-score = 1.89). Within this pathway, 14.6% of the genes (*CCND3, E2F7, E2F8, FHIT, HDAC6, MAP2K2, PIK3CD, PIK3R6, RALB, RASSF1, RASSF5, SIN3A, TFDP1*, *TGFA*) were dysregulated in response to smoky coal combustion.

Several oxidative stress response pathways were also enriched, including glutaminergic receptor signaling, CREB signaling, G-protein coupled receptor signaling, RHO GTPase cycle, and mitochondrial dysfunction. The two inhibited pathways were ‘epithelial adherens junction signaling’, which is critical for maintaining cell-cell adhesion and epithelial barrier integrity, and ‘mitochondrial dysfunction’, which may represent an adaptive response to restore mitochondrial function and mitigate oxidative stress.

#### 3.5.3. Molecular network analysis

The highest-ranked network, associated with “cancer, organismal injury and abnormalities, and respiratory diseases,” had a score of 52 and included 34 molecules, including *ADAMTS8, BANP, CFAP45, CIAO2A, CLDN2, CPEB4, DNAAF5, KHDRBS3, MEK, PLSCR4, RBM20, RBMS1,* and *YTHDF1* (Table S8). The second-highest ranked network, with a score of 50, was related to “cellular development, cellular growth and proliferation, and protein trafficking” and consisted of 33 molecules, including *ANKRD12, CD109, DAPK1, DPP8, E3/ISG15, GRK6, HDAC6, NXNL2, PRDX2, RAD23B, RBM27, S100A6, SLC25A18,* and *XIAP*. These findings underscore key molecular interactions and their potential roles in smoky coal-related biological processes and disease pathways.

## DISCUSSION

This study provides novel insights into the DNA methylation alterations associated with HAP exposure from smoky coal combustion in never-smoking Chinese women in Xuanwei, a population with exceptionally high lung cancer rates ^4,5^. We identified several differentially methylated CpG sites, predominantly exhibiting hypomethylation, in response to HAP exposure. While some smoking-related DNAm signatures, such as *AHRR (*cg05575921) hypomethylation, overlapped with HAP exposure, the majority remained distinct. A life-course assessment identified persistent epigenetic signatures across childhood and cumulative exposures, suggesting early-life PAH exposure may have long-lasting effects, potentially compounded by continued exposure over time. Within the broader PAH clusters, the methylated PAH, 5-MC appears to be a significant contributor to the observed DNAm variations. Integrative bioinformatics analysis revealed activation of cancer-related, neurodevelopmental, immune-regulatory, and G-protein coupled signaling pathways in response to HAP exposure, alongside molecular networks linked to cancer, organismal injury, and major disease pathways.

To our knowledge, this is the first population-based EWAS examining HAP exposure from solid fuel use. A previous population-based study of never-smoking women in Warsaw, Poland (n = 42) reported global hypomethylation associated with both early-life and adulthood exposure to indoor air pollution from solid fuel use but did not assess individual CpG site associations ^32^. In this population, we previously found that PAH exposure, among which 5-MC, was associated with epigenetic age acceleration as measured by the GrimAge clock ^11^, a DNAm-based biomarker linked to mortality and morbidity across various diseases, including cancer ^12^. Additionally, we identified that GrimAge acceleration was prospectively associated with increased future lung cancer risk in never-smoking men and women in Shanghai, China^13^. These findings lay the groundwork for understanding HAP-induced epigenetic modifications and their potential impact on disease risk.

Although the epigenetic changes in response to HAP exposure were largely distinct from smoking-related DNAm patterns, some similarities were observed. At the individual CpG level, some CpG sites previously linked to smoking were also associated with HAP exposure or its PAH constituents. Notably, hypomethylation at cg05575921 (*AHRR*), a well-established smoking-related CpG site, was significantly associated with current exposure to 5-MC, supporting the robustness of our findings despite the limited sample size. Additionally, a modest correlation between cumulative PAH exposure and DNAm-based smoking pack-years (ρ = 0.31, P = 5.1 × 10□□; Table S2) in this never-smoking population suggests that long-term exposure to PAHs from non-smoking environmental sources, such as smoky coal combustion, may contribute to epigenetic signatures that overlap with those typically associated with smoking. Previously identified smoking-related CpG sites that were also linked to smoky coal combustion – annotated to genes such as *AHRR, SNORD93, NWD1, EDC3, STK24, ZDHHC14, HIF3A* (Table S3) – converge on key pathways involved in pollutant response, respiratory diseases, and carcinogenesis. Notably, several of these CpG sites (*SNORD93, NWD1, EDC3*) have been associated with COPD ^33^, which is an established independent risk factor for lung cancer in both smokers and never smokers ^28,29^. DNAm patterns associated with smoky coal exposure may reflect underlying molecular mechanisms common to both COPD and lung cancer, including chronic inflammation and oxidative stress ^34^.

The *AHRR* (aryl hydrocarbon receptor repressor) gene plays a key role in mediating responses to PAHs via aryl hydrocarbon receptor pathway, while the *HIF3A* (hypoxia inducible factor 3α) gene is involved in hypoxia signaling – both pathways relevant to air pollution exposure and tumor microenvironments ^35-38^. In addition, regulatory RNAs such as SNORD93 (small nuclear RNA, C/D box 93) and RNA processing factors such as EDC3 (enhancer of mRNA decapping 3) may modulate gene expression related to cell proliferation ^39,40^. The *NWD1* (NATCH and WD repeat domain containing 1) gene, located on chromosome 19p13.11, has been implicated in hormone-related cancers (ovarian, breast, prostate) and autism spectrum disorder, suggesting a role in tumorigenesis and neurodevelopment ^41-44^. The *STK24* (serine/threonine kinase 24) gene, which regulates MAPK signaling and oxidative stress-induced apoptosis ^30^, has been linked to cell proliferation, migration, and immune modulation in lung adenocarcinoma ^45^. Furthermore, the *ZDHHC14* gene encodes a palmitoyl transferase, which is involved in protein lipidation, cell differentiation and apoptosis ^30,46^.

Our results indicated a strong correlation between DNAm patterns associated with broader PAH clusters and those of 5-MC. Several CpG sites significantly associated with PAH clusters were also linked to 5-MC across exposure windows (Tables S4 and S6). In particular, the effect sizes for PAH clusters and 5-MC at the top epigenome-wide significant CpG sites (782 sites, ^13^ < 0.05; Table S6) were highly correlated (*r* = 0.97; Figure S2). By comparison, DNAm patterns at known smoking-related CpG sites exhibited a more complex interaction with PAH constituents, with 5-MC contributing significantly but not exclusively (*r* = 0.87). These data demonstrate a robust relationship between DNAm variations linked to PAH clusters and 5-MC. However, while the high correlation suggests that 5-MC may play an important role within the broader PAH cluster in driving these DNAm variations, further studies are necessary to determine whether 5-MC is a causal factor or merely a marker of these epigenetic responses.

Our study population, characterized by limited socioeconomic resources and low levels of formal education (77.4% reliance on smoky coal and 67.9% with limited education; Table 1), reflects conditions that contribute to sustained HAP exposure. In Xuanwei, households historically relied on smoky coal for cooking and heating, with many continuing this practice into adulthood, leading to a stable lifetime exposure pattern. This is supported by the strong correlation between childhood and cumulative PAH exposures (ρ = 0.77, P < 2.2 × 10□¹□; Table S2). The consistent associations observed at specific CpG sites (eg20059928, cg09938479, cg04320476 and cg15090217 (*SCL34A2*), cg23209721; Figure 2c and 3c, Table S3 and S6) with both childhood and cumulative exposures suggests that these epigenetic modifications are established early and either persist or further modified by continued exposure over time. We previously found that early-life PAH exposure plays a critical role in determining lung cancer risk in this population ^9^. Here, we provide further evidence that early-life PAH exposure is associated with persistent DNAm variations, which may serve as mechanistic links and early biomarkers for the increased lung cancer risk observed in this population.

Among CpG sites showing consistent association patterns between childhood and cumulative exposures, three (cg04320476, cg15090217, cg12564034) are annotated to the *SLC43A2* gene (Table S6). *SLC43A2* encodes a transporter for neutral amino acids, including methionine, and is expressed in blood, leukocytes, and lung tissues ^30^. Dysregulation of *SLC43A2* has been implicated in tumor-mediated immune evasion, potentially by limiting methionine uptake in CD8 T cells ^47^. Consistent with this mechanism, our cell-type-specific functional overlap analysis using eFORGE (Figure 4) revealed that smoky coal combustion predominantly alters DNAm at T cell regulatory elements, with the strongest enrichment observed in primary T cells (E034 cells; Figure 4). Further analysis revealed that these DNAm alterations were particularly enriched in promoters (H3K4me3), enhancers (H3K4me1), and active transcription start sites (TssA) in CD8 memory T cells. Moreover, our findings align with genome-wide association studies (GWAS), indicating that top lung cancer-associated variants are enriched in E034: primary T cells from peripheral blood, suggesting a potential functional overlap between lung cancer GWAS signals and smoky coal-associated EWAS findings.

Canonical pathway analysis revealed significant activation of cancer-related pathways in response to HAP exposure, including ‘Molecular Mechanisms of Cancer’ and ‘Non-Small Cell Lung Cancer Signaling’ (Figure 5, Table S7). Additionally, pathways associated with neurodevelopment and synaptic function (e.g., autism signaling, CREB signaling in neurons, netrin signaling), inflammation and immune regulation (e.g., phagosome formation, estrogen receptor signaling), and G-protein-coupled receptor and signal transduction (e.g., G-protein coupled receptor signaling, G alpha (q) signaling, S100 family signaling, RHO GTPase cycle) were significantly enriched. Consistent with our previous findings, G-protein coupled receptor signaling pathways were also enriched in oral cell DNAm patterns linked to lung cancer development among never-smokers in the prospective Shanghai cohorts ^13^.

In contrast, two pathways were significantly inhibited in response to smoky coal combustion. The suppression of the mitochondrial dysfunction pathway suggests a compensatory cellular response to restore homeostasis, while the inhibition of epithelial adherens junction signaling may reflect a compromised epithelial barrier, potentially increasing susceptibility to environmental toxins and inflammation. Additionally, IPA analysis identified molecular networks associated with cancer, organismal injury and abnormalities, cell growth and proliferation, and respiratory, neurological, and cardiovascular disorders (Table S8).

Our study has several notable strengths. First, it exclusively focused on never-smoking Chinese women in a region with one of the highest lung cancer rates worldwide ^5^, eliminating potential confounding factors related to gender, race/ethnicity, and cigarette smoking. We implemented rigorous personal air monitoring among study participants, enabling accurate exposure assessment through robust statistical approaches. Additionally, the life-course approach allowed us to examine the effects of PAH constituents across childhood, current, and cumulative exposure windows, providing insights into both short- and long-term impacts on leukocyte DNA methylation patterns. However, our analysis utilized a list of smoking-related CpG sites identified in populations of European ancestry, which may not fully capture epigenetic variations in the Chinese population. Furthermore, the relatively small sample size may have limited statistical power to detect associations with smaller effect sizes. Given the high carcinogenicity of smoky coal, we initially anticipated significant biological effects on biomarkers associated with cancer development and mortality.

## CONCLUSIONS

This study highlights distinct epigenetic alterations associated with HAP exposure from smoky coal combustion in never-smoking Chinese women, a population with high lung cancer rates. Differentially methylated CpG sites, primarily exhibiting hypomethylation, were identified, with some overlap but largely distinct patterns from smoking-related DNAm signatures. Life-course analysis suggests that early-life PAH exposure may have lasting impacts on DNAm patterns. Within the broader PAH clusters, the methylated PAH, 5-MC appears to be a significant contributor to the observed DNAm variations. Bioinformatics analysis further revealed activation of pathways involved in carcinogenesis, immune regulation, neurodevelopment, and signal transduction, underscoring the potential long-term health impacts of HAP exposure. Future studies with larger sample sizes are needed to validate these findings.

## Data Availability

All data generated or analyzed during this study are included in this published article and its supplementary information files.

## LIST OF ABBREVIATIONS

AHR: Aryl Hydrocarbon Receptor
AHRR: Aryl Hydrocarbon Receptor Repressor
BC: Black Carbon
BMI: Body Mass Index
BMIQ: Beta Mixture Quantile Dilation
CD8: Cluster of Differentiation 8 Positive
COPD: Chronic Obstructive Pulmonary Disease
CpG: Cytosine-phosphate-Guanine
CREB: cAMP Response Element Binding
DNAm: DNA Methylation
DNN: Deep Neural Network
EWAS: Epigenome-Wide Association Study
FDR: False Discovery Rate
GEE: Generalized Estimating Equations
GWAS: Genome-Wide Association Study
HAP: Household Air Pollution
HIF3A: Hypoxia Inducible Factor 3 Alpha
IPA: Ingenuity Pathway Analysis
NO: Nitrogen Dioxide
NSCLC: Non-Small Cell Lung Cancer
PAH: Polycyclic Aromatic Hydrocarbon
PAH31: 31 Polycyclic Aromatic Hydrocarbon Constituents (Current Exposure)
PAH33: 33 Polycyclic Aromatic Hydrocarbon Constituents (Childhood Exposure)
PAH36: 36 Polycyclic Aromatic Hydrocarbon Constituents (Cumulative Exposure)
PM: Particulate Matter
RHO: Ras Homolog Gene Family
RNA: Ribonucleic Acid
SLC43A2: Solute Carrier Family 43 Member 2
SNP: Single Nucleotide Polymorphism
SO: Sulfur Dioxide
TSS: Transcription Start Site
ZDHHC14: Zinc Finger DHHC-Type Palmitoyl transferase 14
5-MC: 5-Methylchrysene

## Consent for publication

Not applicable

## Availability of data and materials

All data generated or analyzed during this study are included in this published article and its supplementary information files. For original data, please contact the corresponding author, Mohammad L. Rahman at

## Authors’ contributions

N.R., Q.L., R.V. and Y.H. designed the study and supervised data collection. M.L.R. prepared the analysis plan, analyzed the data, and drafted the manuscript. All authors contributed to the interpretation of the results and revision of the manuscript for important intellectual content and approved the final version of the manuscript. M.L.R. and Q.L. are the guarantors of this work and, as such, had full access to all the data in the study and take responsibility for the integrity of the data.

## Acknowledgements

The authors acknowledge the research contributions of the Cancer Genomics Research Laboratory of the Intramural Research Program, National Cancer Institute, National Institutes of Health for their expertise, execution, and support of this research in the areas of project planning, wet laboratory processing of specimens, and generating the data.

