## Supplemental Figures for "Epigenome-wide association study of household air pollution exposure in an area with high lung cancer incidence"

**Figure S1**. Beta density plots of leukocyte DNA methylation in never-smoking

women. The x-axis shows the DNA methylation beta values, while the y-axis

indicates their density. Panel A shows the plot before data quality control, Panel B

after quality control, and Panel C after normalization using the beta mixture quantile

dilation (BMIQ) method.

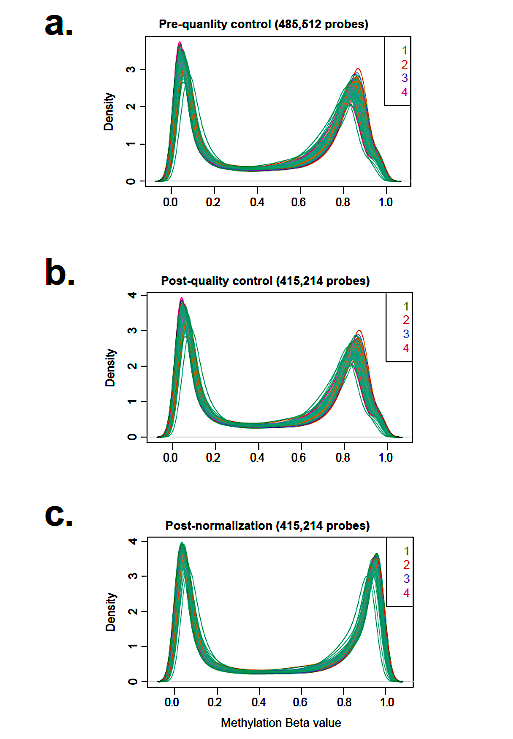

**Figure S2**. Pearson’s correlations between beta coefficients of PAH clusters (x-axis) and 5-

MC (y-axis) across respective exposure windows for (a) previously identified smoking-related

CpG sites (2,476 CpGs) and (b) top epigenome-wide significant sites (782 CpGs; FDR < 0.05).

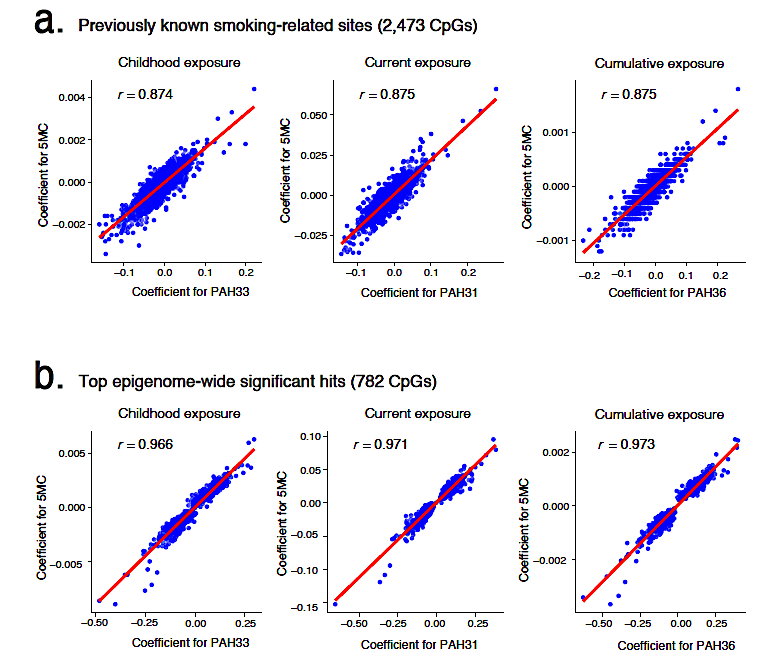
